# Speech Intelligibility Index (SII) as a Potential Referral Metric for Adult Cochlear Implant Candidacy Evaluation

**DOI:** 10.64898/2026.09.10.26362790

**Authors:** Jennifer A. Kong, Terrin N. Tamati, Aaron C. Moberly, Jonathan D. Neukam

## Abstract

**Objectives:** The purpose of this pilot study was to investigate (1) if the Speech Intelligibility Index (SII) could be used to identify ears that meet cochlear implant (CI) candidacy beyond the “60/60” referral guidelines and (2) investigate the relationship between SII from patients’ own hearing aids (HAs) and SII from optimally fit clinic-stock HAs such that a patient’s own HAs can act as a proxy for an optimally fit HA and thus as a referral metric.

**Design:** Prospective study of 23 participants (≥18 years) with bilateral moderate to profound sensorineural hearing loss who were CI candidates in at least one ear. SII-Clinic (SII-C) values were recorded with probe-microphone measures using a Verifit II device (NAL-N2 targets) with speech stimuli presented at 60 dB A during patient CI evaluation in either their own HAs or clinic-stock HAs. SII values from participants’ own HAs were recorded as part of a larger research protocol, thus SII-Personal (SII-P), with probe-microphone measures using a Verifit I device (NAL-RP targets) with pink noise stimuli presented at 65 dB A. Participants that did not wear HAs did not have SII-P calculated. Data was collapsed across ears. Correlation analysis between SII and best-aided consonant–nucleus–consonant (CNC) was conducted. Benchmark SII values were extrapolated using a best-fit line and CI candidacy rates reflecting various aided CNC scores that could be applied across CI centers. Nested logistic regression models were used to determine if the SII-C could improve model fit beyond the revised ear-specific “60/60” referral guideline. The relationship between SII-C and SII-P was assessed with Wilcoxon signed-rank test and Spearman correlation.

**Results:** Correlation between SII-C and aided CNC score was r = 0.82 (p < 0.001), and the correlation between SII-P and aided CNC was r = 0.46 (p = 0.014). Extrapolated benchmarks of SII were calculated based on different CNC candidacy cutoffs, and benchmarks were able to capture at least 83% of the ears that met CNC candidacy cutoffs. SII was also found to improve model fit with word recognition score (WRS) and pure tone average (PTA) for 60% CNC candidacy cutoff (χ ^2^(1) = 9.7; p = 0.002) and increased discrimination (AUC = 0.89 to AUC = 0.99). There was no significant difference between SII-C and SII-P using the Wilcoxon signed-rank test (p = 0.559).

**Conclusions:** A larger study is needed that includes more ears that do not qualify for a CI; however, the results of this study demonstrate that (1) SII has the potential to be used in conjunction with the “60/60” referral guideline for CI candidacy evaluations and (2) SII can be used as a marker of audibility across different devices, including patients’ own HAs, making SII an accessible metric during HA fittings and fine tunings.

## Introduction

A cochlear implant (CI) is a neural prosthetic used as the standard-of-care for treatment of individuals with moderate to profound levels of hearing loss, a condition associated with significant medical and psychosocial consequences, including mental illness, loneliness, and neurocognitive dysfunction (Lin et al. 2013; Maharani et al. 2019; Wilson et al. 2017). CIs provide access to sound which can improve speech understanding leading to better quality of life including communication and social participation (McRackan et al. 2018a; McRackan et al. 2018b; Kelsall et al. 2021; Gaylor et al. 2013; Buchman et al. 2020). Despite the established benefits, CI utilization in the United States remains low, with an estimated 2% to 13% of eligible adults receiving a device (Nassiri et al. 2022b). Some known barriers to CI uptake are lack of referral from non-CI professionals (Mashal et al. 2022; Reddy et al. 2022) and inconsistent referral criteria (Nassiri et al. 2022a), including inadequate testing materials available to accurately assess unaided speech perception by referring providers (Greiner et al. 2023). Current referral recommendations in the United States follow the “60/60” referral guideline which uses set criteria of speech perception scores and puretone average (PTA), however, exploration of other clinically available nonbehavioral referral metrics, such as the speech intelligibility index (SII), has not been explored in adults.

### Referral Guidelines for Cochlear Implant Candidacy Evaluation – Unaided Testing

Initial referral guidelines typically utilize unaided audiological testing metrics to predict CI candidacy. For example, routine unaided monosyllabic word recognition scores (WRS) and pure tone average (PTA; 500, 1000, and 2000 Hz) can be used with relatively high sensitivity and specificity to identify patients that would qualify for a CI (Gubbels et al. 2017; Hunter & Tolisano 2021). The use of unaided audiological measures led to development and validation of the well-known “60/60” referral guideline, which recommends referral for a CI candidacy evaluation when the better hearing ear has a PTA ≥60 dB HL and unaided WRS in quiet ≤60% correct (Zwolan et al. 2020). However, the “60/60” referral guideline was developed to identify traditional CI candidates (e.g., those who would qualify for a CI in both ears), which aligned with the Minimum Speech Test Battery (MSTB) at that time. This former version of the MSTB focused on pre-and post-CI assessments in traditional CI candidates, and the associated MSTB document provided no clear recommendations on candidacy or referral (Advanced Bionics et al. 2011). Because of candidacy expansion that extends beyond traditional candidates, the MSTB Version 3 (MSTB-3; Dunn et al., 2024) now recommends CI candidacy, and therefore referral, using ear-specific testing criteria. This change was motivated by research demonstrating benefits of cochlear implantation in non-traditional candidates (e.g., single-sided deafness [SSD], asymmetric hearing losses [AHL], and those who may benefit from electric-acoustic stimulation [EAS]). As a result, the MSTB-3 makes recommendations for CI referral based on the individual ear being considered for implantation, rather than the performance of the better-hearing ear or both ears of the patient. Accordingly, the “60/60” referral guideline has been revised to recommend CI candidacy evaluation referral for a patient with a PTA ≥60 dB HL and unaided WRS in quiet ≤60% correct “in the ear to be implanted” (i.e., typically the patient’s worse ear) (Dunn et al. 2024).

Although this revised ear-specific “60/60” referral guideline proposed by the MSTB-3 has the advantage of using routine unaided audiological measures of WRS and PTA, there are limitations to this approach. First, as with any screening tool, the “60/60” referral guideline is not perfectly sensitive nor specific (Lee et al. 2022). Second, unaided monosyllabic word recognition testing (WRS) is sometimes performed using live-voice stimulus presentation, despite AAA/ASHA guidelines that recommend recorded materials as the preferred method (American Speech-Language-Hearing Association n.d.; Hornsby & Mueller 2013). Additionally, presentation level may be insufficient resulting in reduced WRS scores. As a result, WRS may not be accurate and, if used as a CI referral criterion, may lead to inappropriate referrals or non-referrals. Third, unaided PTA across 500, 1000, and 2000 Hz provides only a rough estimate of hearing status and provides no information about the configuration of a patient’s hearing loss. In other words, a patient with a mild-sloping-to-profound hearing loss can have the same unaided PTA as a patient with a flat severe hearing loss, but these two patients may respond very differently to amplification and challenging listening environments. Thus, additional or complimentary referral criteria beyond the “60/60” referral guideline could be helpful in identifying patients who are appropriate for referral for CI evaluation.

### The Cochlear Implant Candidacy Evaluation – Best-aided Testing

At the CI candidacy evaluation following referral, the MSTB-3 prioritizes the use of best-aided individual-ear consonant–nucleus–consonant (CNC) words (Peterson & Lehiste 1962) as the primary speech recognition test for determining CI candidacy. Prior versions of the MSTB (Advanced Bionics et al. 2011; Nilsson et al. 1996) recommended a combination of aided CNC word recognition with aided AzBio and BKB-SIN sentence recognition scores. This shift in candidacy determination was based on a change in FDA-approved indications for EAS, SSD, and AHL CI candidates, given the lower likelihood of post-operative ceiling effects with words than with sentence materials (Gifford et al. 2008; Sladen et al. 2017). Notably, the MSTB-3 does not set specific CNC candidacy guidelines but rather allows each CI center to establish its own CI candidacy cutoffs based on best-aided CNC scores (e.g., ≤60-, ≤50-, or ≤40-% correct), with cutoffs that “should align with the clinician and the center seeing the patient.”

An essential aspect of the CI evaluation is testing each ear in its “best-aided” listening configuration, using a HA that has been optimized for the hearing loss in that individual ear. The MSTB-3 recommends real-ear verification of the patient’s HA output via probe-microphone measurement at the time of the CI candidacy evaluation to ensure that the gain and output of the HA match the patient’s ear-specific hearing loss using evidence-based prescriptive targets (Mueller & Picou 2010). If the patient’s own HAs do not meet prescriptive targets, it is recommended that clinic-stock HAs be programmed, verified, and used during aided testing, thus reflecting ear-specific “best-aided” performance.

### Speech Intelligibility Index (SII) as a Potential Referral Metric for Cochlear Implant Candidacy Evaluation – The Current Study

Already in use for pediatric CI referral (Holder et al. 2026; McCreery 2014; Stiles et al. 2012; Wiseman et al. 2023), the Speech Intelligibility Index (SII; ANSI S3.5-1997 R2020) is a promising tool for adult CI referral. The aided SII is used to characterize the aided access to speech information provided by the HA, with 0.0 representing no audible speech and 1.0 representing full audibility. Unlike more complex batteries, this metric can be obtained during routine HA fittings and fine tunings. Therefore, this pilot study aimed to evaluate the SII as a supplemental referral metric to the “60/60” referral guideline by establishing the relationship between SII and adult CI candidacy using patients that qualified for a CI in at least one ear and underwent CI surgery. Our first hypothesis was that the SII acquired with optimally fit HAs could identify ears that meet CI candidacy and provide additional information above and beyond the “60/60” guideline determined by incremental logistic regression modeling and ROC analysis.

The utility of SII as a CI referral criterion depends on the degree to which the patients’ own HAs are optimized to their hearing loss and whether an SII recorded from the patient’s own device could be used as a referral metric. Patients undergoing CI candidacy evaluation are often underfit with their own HAs due to lack of verification measures at HA fittings or follow-up and/or transducer and coupling limitations. Additionally, HAs may not always be programmed to prescriptive targets due to patient preference or comfort. Because of this fact, clinic-stock HAs are often required to conduct “best-aided” testing during the CI evaluation (Prentiss et al. 2020); however, differences between clinic-stock HAs and the patients’ own HAs has been poorly defined. Thus, the secondary purpose of the current study was to quantify the degree to which the SII for patients’ own HAs differs from the SII of a best-aided clinic HA. We hypothesized that the SII from patients’ own HAs would be significantly lower than the SII acquired with optimally fit clinic-stock HAs, but that the two SIIs would be strongly correlated. A strong correlation between these two SII measures would provide support that even a suboptimal SII of patients’ own HAs could serve as a useful real-world indicator for CI referral.

## Materials and Methods

### Participants

Participant inclusion included adult CI candidates (≥18 years) with bilateral moderate to profound sensorineural hearing loss and best-aided CNC speech recognition ≤60% words correct in the ear to-be-implanted. Bilateral moderate to profound hearing loss was defined as having at least two adjacent puretone thresholds ≥ 45 dB HL in both ears. This criteria includes asymmetrical hearing loss while excluding single-sided deafness. Best-aided condition refers to guidelines established by the MSTB-3 which involves speech testing presented at 60 dB A in soundfield with a HA that has been optimized for the hearing loss in an individual ear. For the remainder of this study, best-aided will be referred to as “aided CNC”.

Participants were recruited following their CI evaluation appointment, via in-person recruitment by lab members. Following informed written consent, clinical data were collected and recorded from patient electronic medical records. A separate in-lab research session was scheduled prior to implantation (typically within one month of the CI evaluation appointment), during which SII metrics were collected and recorded from participants’ personal hearing devices (referred to as SII-P).

Aided SII values from the clinical CI evaluation (SII-C) were obtained with probe-microphone measures on a Verifit II (Audioscan, Ontario, CA) using 60 dB A speech stimuli and NAL-N2 targets. If the patient’s own HAs met target, that SII was recorded as the SII-C; otherwise, a clinic-stock HA (Resound Enzo or Phonak Naida) was fit with a foam earplug, and that SII-C was recorded. Insertion gain was calculated from the manufacturer’s average unaided response.

For in-lab testing, the SII-P was obtained with probe-microphone measures on Verifit I using 65 dB A pink noise and NAL-RP targets. SII-P was not recorded for participants who did not wear HAs which was either due to lack of ownership of HAs or the wearing of other personal hearing devices not classified as HAs (e.g., Pocket Talker). Insertion gain was calculated from each participant’s unaided response. The discrepancy between verification (SII-C versus SII-P) measures was due to the CI candidacy evaluation using 60 dB A speech stimuli for speech testing while the research session used 65 dB A speech stimuli for speech testing (research-based speech results not reported in this study). Average insertion gain values during the clinic visit were used due to time limitations of the clinical appointment while the use of pink noise, rather than speech stimuli, during the research session was also due to time limitations.

This study was conducted under Institutional Review Board 221985. Informed written consent was obtained from all participants.

### Analysis

All analyses were performed in MatLab (Natick, MA, USA). Unaided WRS testing, aided CNC testing, and SII distributions were described using descriptive statistics. Due to the current focus on ear-specific assessments, ear-specific analyses were carried out. Data was collapsed across the ear-to-be-implanted and the contralateral ears due to the small sample size and the pilot nature of this study, thus, collapsing across ears permitted inclusion of ears with a range of baseline PTA, unaided WRS, aided CNC, and SII.

To test the first hypothesis, that the SII could contribute to CI “60/60” referral guidelines, three analyses were performed. First, correlation analyses were performed for both SII-C and SII-P with aided CNC scores from the CI candidacy evaluation. Second, benchmark SII-C and SII-P values were extrapolated using a best-fit line and CI candidacy rates reflecting various aided CNC scores that could be applied across CI centers based on MSTB-3 suggestions (Dunn et al. 2024). The extrapolated SII-C or-R value could be considered as a benchmark in identifying patients who would meet candidacy (e.g., Gubbels et al. 2017).

Lastly, multiple logistic regression models were fit to predict CI candidacy (as determined by CNC cutoffs of CNC ≤60%, ≤50%, or ≤40% words correct) using referral measures including (1) WRS and PTA and (2) WRS, PTA, and SII-C as predictors. Each CI center must establish their own CI candidacy guidelines; therefore, in order to generalize across CI centers, analyses were performed for varying degrees of CI candidacy stringency (i.e., aided CNC ≤60%, ≤50%, or ≤40% words correct). Nested models with increasing predictor sets were compared to evaluate the incremental contribution of SII-C to the prediction model of who and who is not a CI candidate based on aided CNC. Receiver Operator Characteristic (ROC) curve analysis was used to quantify and compare model discrimination using Area Under the Curve (AUC) derived from predicted probabilities of the logistic regression models. As a sensitivity analysis, mixed-effects logistic regression models with subject-level random intercepts were fit. Because these models yielded findings similar to the primary logistic regression analyses for the retained cutoffs, and because some models were unstable at the 60% cutoff (i.e., too many CI candidates compared to the overall N), the standard logistic regression results are presented in the main text with mixed-effects modeling presented in supplementary materials.

To test the second hypothesis, that the mean SII-C would be significantly higher (a < 0.05) than the mean SII-P, a Shapiro-Wilks test was first used to test for normality followed by a Wilcoxon Signed-Rank test. Spearman correlation was used to describe the relationship between SII-C and SII-P collapsed across ears.

## Results

Twenty-three participants (8 female/15 male; N = 46 ears) were enrolled with a mean age of 70.6 years (SD 10.2). Thirteen wore bilateral behind-the-ear HAs, and a single participant wore a single in-the-ear HA. Six did not wear HAs and three used a Pocket Talker. SII-C was calculated by patients’ own HAs in five patients and by clinic-stock HAs in 18 patients. For SII-P, participants who did not wear HAs were excluded, leading to an N = 14 participants or 28 ears for analyses including SII-P. Unaided WRS was unavailable for both ears in one participant and in the contralateral ear of a subsequent participant leading to a reduced N of 43 ears with available WRS.

The overall (N = 43) mean unaided WRS was 31.3% (SD 24.5). The overall (N = 46) mean PTA of 77.2 dB HL (SD 16.0). The mean aided CNC score was 77.2% (SD 26.8) and the mean sentences in quiet score (AzBioQ) was 43.2% (SD 33.5). The overall mean SII-C was 30.0 (SD 15.5; N = 46) and the mean SII-P was 28.4 (SD 16.8; N = 28). Forty-one of the 46 ears had a PTA greater than 60 dB HL and 38 had a WRS score less than 60%. Forty ears had aided CNC scores ≤60% (40/46). Thirty-three ears had aided CNC scores ≤50% (33/46). Twenty-nine ears had aided CNC scores ≤40% (29/46). See Table 1 for a summary of participant characteristics.

**Table 1:** Participant characteristics.

|  | N | All ears<br>Mean (SD)<br>(MIN–<br>MAX) | N | Ear to-be-implanted Mean<br>(SD)<br>(MIN–MAX) | N | Contralateral<br>ear<br>Mean (SD)<br>(MIN–MAX) |
| --- | --- | --- | --- | --- | --- | --- |
| Unaided WRS (%) | 43 | 31.3 (24.5)<br>(0–90) | 22 | 19.7 (16.5)<br>(0–60) | 21 | 42.9 (26.0)<br>(0–90) |
| Puretone average<br>(dB HL) | 46 | 77.2 (16.0)<br>(45–120) | 23 | 80.1 (14.7)<br>(55–120) | 23 | 74.4 (17.0)<br>(45–113) |
| Aided CNC (%) | 46 | 33.1 (26.8)<br>(0–94) | 23 | 26.52 (20.3)<br>(0–60) | 23 | 40.0(31.0)<br>(0–94) |
| Aided AzBioQ (%) | 46 | 43.2 (33.5)<br>(0–97) | 23 | 36.9 (30.7)<br>(0–89) | 23 | 49.6 (35.6)<br>(0–97) |
| SII from clinic (SII-C) | 46 | 30.0 (15.5)<br>(0–62) | 23 | 26.2 (12.5)<br>(0–47) | 23 | 33.8 (15.7)<br>(3–62) |
| SII from research<br>(SII-P) | 28 | 28.4 (16.8)<br>(0–59) | 14 | 23.5 (14.1)<br>(0–40) | 14 | 33.3 (18.3)<br>(0–59) |

WRS = word recognition score, PTA = pure tone average, CNC = consonant–nucleus–consonant, AzBioQ = AzBio sentence test in quiet, SII-C = Speech Intelligibility Index from clinic, SII-P = Speech Intelligibility Index from Personal/Research

### Analysis I – Relating SII to CI candidacy

The correlation between SII-C and aided CNC score was r = 0.82 (p < 0.001), and the correlation between SII-P and aided CNC was r = 0.46 (p = 0.014). Extrapolated benchmark SII values were calculated by finding the intercept of the aided CNC stringency cutoff and the best-fit linear regression. Forty ears met the CNC ≤ 60% candidacy cutoff and an extrapolated benchmark value of SII-C = 48 was found to capture 100% (40/40) of those who would qualify, and SII-P = 65 was found to capture 100% (25/25). Thirty-three ears met the CNC ≤ 50% candidacy cutoff, and an extrapolated benchmark value of SII-C = 41 was found to capture 94% (31/33), and SII-P = 51 was found to capture 95% (18/19). Twenty-nine ears met the CNC≤ 40% candidacy cutoff, and an extrapolated benchmark value of SII-C = 35 was found to capture 83% (24/29), and SII-P = 36 was found to capture 81% (13/16). See Figure 1.

**Figure 1:**
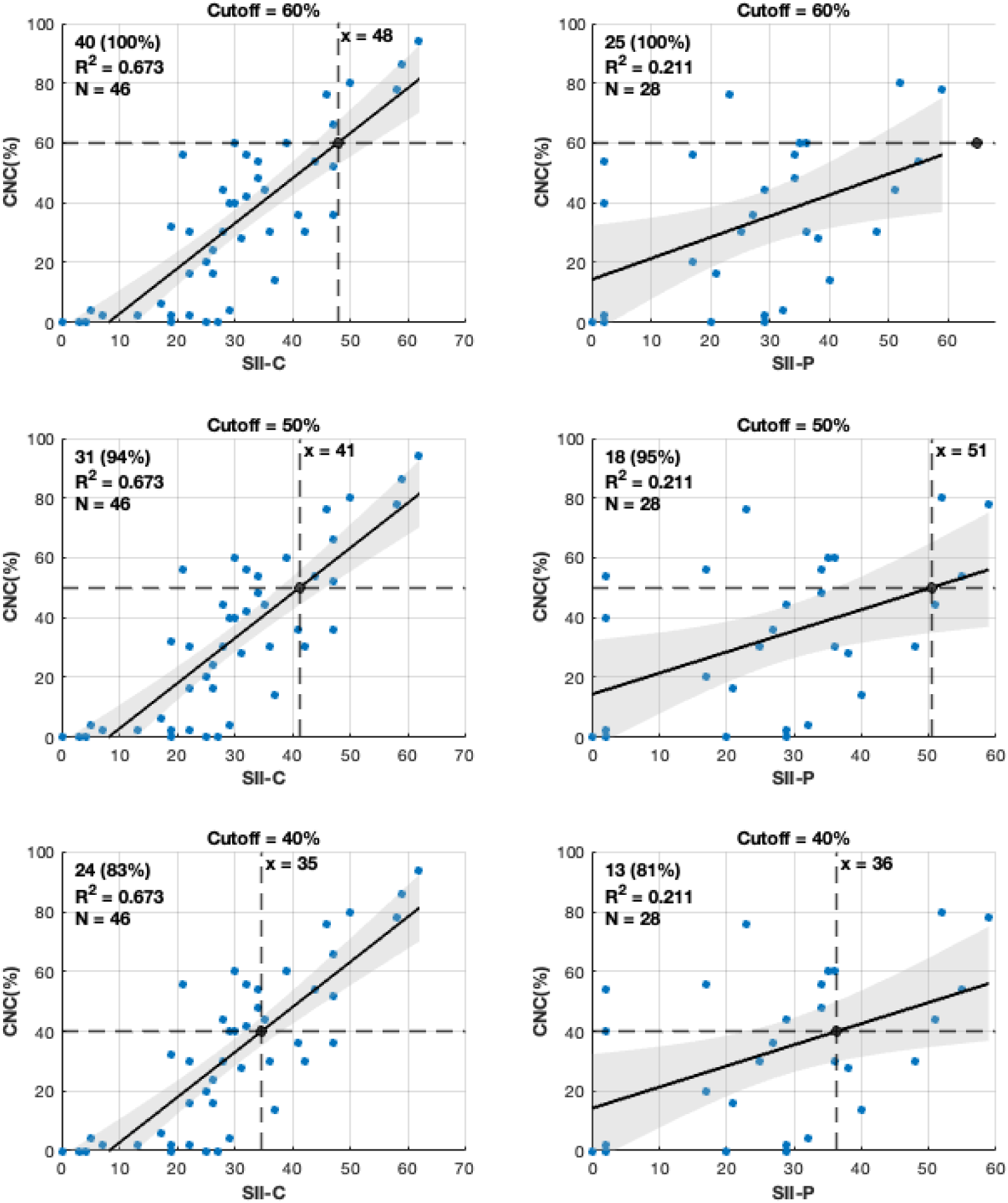
Scatterplot of Speech Intelligibility Index and consonant-nucleus-consonant (CNC) score cutoffs (≤ 60%, 50%, 40%). Best-fit black line shown with 95% confidence interval shaded band. The upper left-hand corner of each plot shows the number of ears captured by the extrapolated benchmark (vertical dashed line) and proportion of total ears that meet CI candidacy at each CNC cutoff (ears falling below the horizontal dashed line). R^2^ values shown to demonstrate the accounted variance. Left-hand column is for SII-C while right-hand column is for SII-P. C.I. = Confidence Interval, CNC = consonant–nucleus–consonant, SII-C = Speech Intelligibility Index from Clinic, SII-P = Speech Intelligibility Index from Personal/Research

Across all three aided CNC candidacy thresholds, the logistic regression models including WRS and PTA showed good discrimination for referral status, and the addition of SII-C significantly improved model fit and discrimination at each cutoff. AUC increased from 0.89 to 0.99 for the 60% criterion, from 0.88 to 0.93 for the 50% criterion, and from 0.82 to 0.88 for the 40% criterion. Full model-fit results for SII-C are shown in Table 2, and corresponding SII-P results are presented in Appendix 1, Supplemental Content. Mixed-effects logistic regression models accounting for subject-level clustering yielded similar results to those of the standard logistic regression for SII-C (see Appendix 2, Supplemental Content).

**Table 2:**
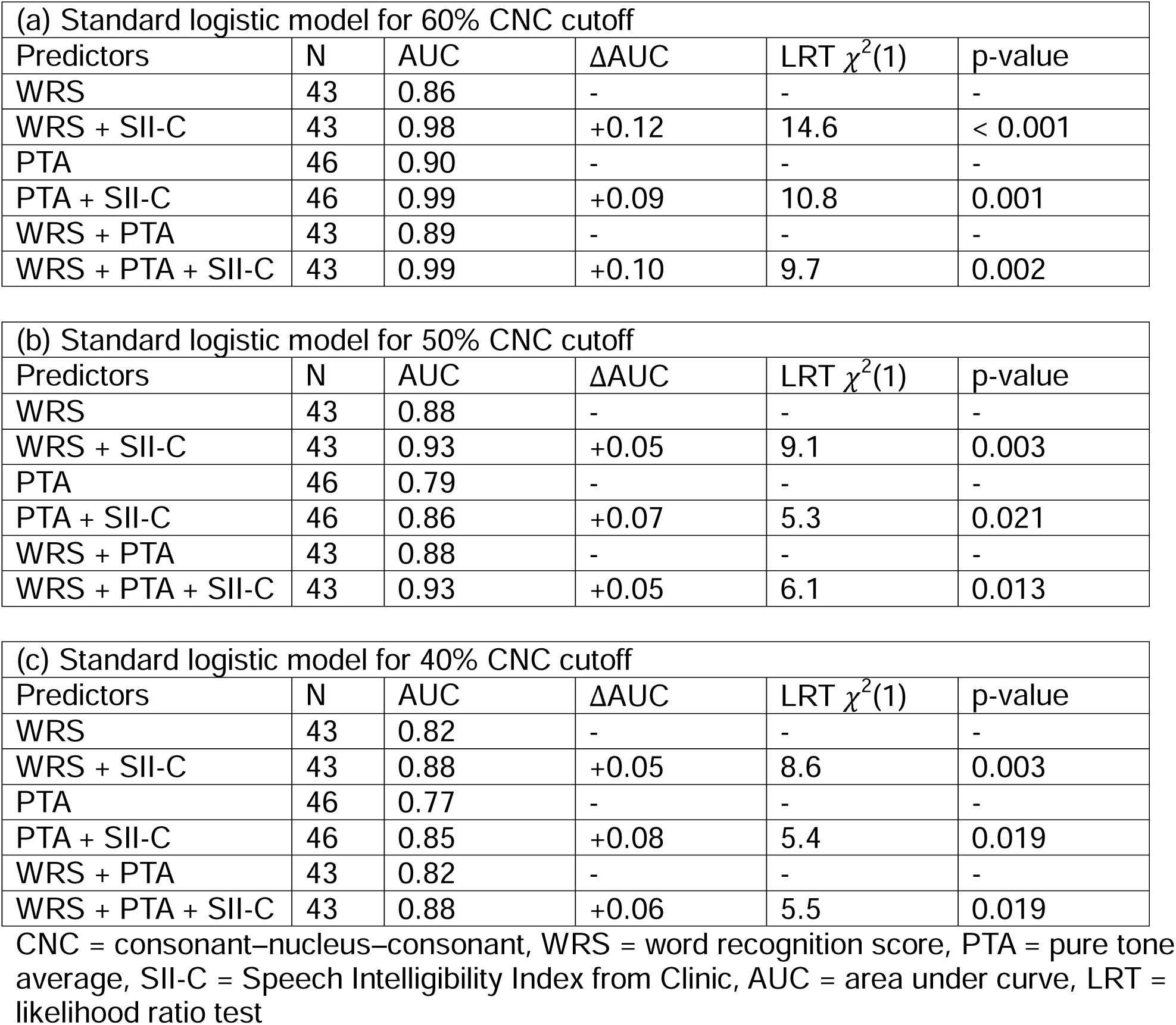
Standard logistic model performance and comparison of area under the receiver operating characteristic curve (AUC) and likelihood ratio test (LRT) results with word recognition scores (WRS), pure tone average (PTA), and Speech Intelligibility Index-C (SII-C) for different consonant-nucleus-consonant (CNC) score candidacy cutoffs: (a) 60% CNC cutoff, (b) 50% CNC cutoff, and (c) 40% CNC cutoff. WRS data were unavailable for three participants, resulting in a reduced sample size (N = 43).

| (a) Standard logistic model for 60% CNC cutoff |  |  |  |  |  |
| --- | --- | --- | --- | --- | --- |
| Predictors | N | AUC | ΔAUC | LRT $\chi^2(1)$ | p-value |
| WRS | 43 | 0.86 | - | - | - |
| WRS + SII-C | 43 | 0.98 | +0.12 | 14.6 | < 0.001 |
| PTA | 46 | 0.90 | - | - | - |
| PTA + SII-C | 46 | 0.99 | +0.09 | 10.8 | 0.001 |
| WRS + PTA | 43 | 0.89 | - | - | - |
| WRS + PTA + SII-C | 43 | 0.99 | +0.10 | 9.7 | 0.002 |

| (b) Standard logistic model for 50% CNC cutoff |  |  |  |  |  |
| --- | --- | --- | --- | --- | --- |
| Predictors | N | AUC | ΔAUC | LRT $\chi^2(1)$ | p-value |
| WRS | 43 | 0.88 | - | - | - |
| WRS + SII-C | 43 | 0.93 | +0.05 | 9.1 | 0.003 |
| PTA | 46 | 0.79 | - | - | - |
| PTA + SII-C | 46 | 0.86 | +0.07 | 5.3 | 0.021 |
| WRS + PTA | 43 | 0.88 | - | - | - |
| WRS + PTA + SII-C | 43 | 0.93 | +0.05 | 6.1 | 0.013 |

| (c) Standard logistic model for 40% CNC cutoff |  |  |  |  |  |
| --- | --- | --- | --- | --- | --- |
| Predictors | N | AUC | ΔAUC | LRT $\chi^2(1)$ | p-value |
| WRS | 43 | 0.82 | - | - | - |
| WRS + SII-C | 43 | 0.88 | +0.05 | 8.6 | 0.003 |
| PTA | 46 | 0.77 | - | - | - |
| PTA + SII-C | 46 | 0.85 | +0.08 | 5.4 | 0.019 |
| WRS + PTA | 43 | 0.82 | - | - | - |
| WRS + PTA + SII-C | 43 | 0.88 | +0.06 | 5.5 | 0.019 |

CNC = consonant–nucleus–consonant, WRS = word recognition score, PTA = pure tone average, SII-C = Speech Intelligibility Index from Clinic, AUC = area under curve, LRT = likelihood ratio test

### Analysis II – Comparing Patient HAs with Optimally Fit HAs using SII

When comparing SII-C to SII-P collapsed across ears and excluding those that did not wear HAs (N = 28 ears), there was no significant difference in means using the Wilcoxon signed-rank test (30.0 [SD 13.1] vs 28.4 [SD 16.8]; W = 199; p = 0.559). The Spearman correlation between the SII-C and SII-P was r = 0.57 (p = 0.002).

## Discussion

The sample size of the current study precludes drawing definitive conclusions or generalizing the findings to the broader clinical population. Nevertheless, the results provide preliminary evidence to support three key concepts: (1) the SII can contribute to the current guidelines or serve as its own benchmark for capturing patients that meet criteria for CI referral across varying levels of aided CNC stringency; (2) for patients who wore HAs in this cohort, the SII values obtained from their own HAs can act as a proxy for the SII of well-fit HAs commonly used in CI evaluations. A larger study is needed to evaluate the effectiveness of the SII as a metric for identifying patients who do not meet CI candidacy given this limited study population consisted of only those who qualified for CI candidacy in at least one ear at the ≤60% CNC cutoff.

It is well established that lack of CI referral is linked to low utilization rates in the United States and worldwide (Greiner et al. 2023; Lamb et al. 2023; Looi et al. 2017; Mashal et al. 2022; Patro et al. 2022; Reddy et al. 2022; Sorkin & Buchman 2016). The development of the “60/60” referral guideline has helped to improve clinical clarity but may still not be a perfect solution for CI referral. The current study findings suggest that in cases where incomplete information is available (WRS or PTA), both the SII recorded from patient’s own devices and from a well-fit stock device is a continuous metric providing nuanced clinical information about patients’ performance level with amplification and the likelihood that they would qualify for and benefit from a CI. Moreover, SII-C demonstrated a statistically significant increase in predictive accuracy. The SII-C was able to capture at least 80% of patients who should undergo a CI evaluation using the most stringent CNC cutoff (≤40%), allowing for applicability despite clinic-specific protocols. This is especially relevant where the SII is lower than an established benchmark (e.g., Similar to the < 0.60 from Holder et al. 2026) or where the SII shows a decline over serial HA fittings. Taking the mean of the three benchmark values form this study, an SII < 0.41 could be used to capture patients that would qualify for a CI.

Furthermore, the results suggest that the SII can act as an additional or complimentary referral metric paired with PTA or WRS or both. There is no established consensus on the frequency of performing pure-tone audiometry in adults (Tsai Do et al. 2024). It is generally left up to the discretion of the audiologist and may depend on factors such as suspected hearing changes or insurance reimbursement. Therefore, when HA patients undergo routine fine tuning or other adjustments, unaided pure-tone audiometry and WRS may not be completed. The SII is a useful metric that is generated from probe-microphone measures and is reported to be used in 68% of audiologists routinely fitting HAs (Jorgensen et al. 2022). With the SII having increased accessibility to patients, it provides the opportunity for more patients to be screened and referred if needed. However, it is important to note that SII is dependent upon accurate measure of puretone thresholds which are also used to calculate PTA.

The term “well-fit” HA in the context of CI evaluation must include the caveat that more appropriate HAs provided by a clinic may nonetheless provide inadequate gain due to transducer limitations and acoustic coupling constraints. Importantly, these shortcomings are still reflected in the SII. Contrary to the secondary hypothesis in the current study that personal HAs would be underfit compared to clinic-stock devices, the mean SII-C was not significantly different than the mean SII-P, and the two SII’s were moderately correlated. Taken together, these findings suggest that, at least for this cohort, the SII remains a valid marker of audibility across different devices.

Lastly, this pilot study warrants the need for a larger study assessing the sensitivity, specificity, and positive and negative predictive values of the SII to screen for CI candidacy, in isolation or in addition to WRS, PTA, and the “60/60” referral guideline. To do so will require a much larger sample of patients, including a large group that does and a large group that does not qualify for a CI. It is noteworthy that the Common Procedural Terminology 2026 (CPT 2026) now includes behavioral verification in soundfield for aided speech perception in quiet and noise (92638), which may encourage more widespread performance of aided speech perception abilities outside of CI centers. It is possible that a rich dataset of aided speech recognition scores (CNC) and SII metrics from patients’ own HAs could be collected to perform such a study.

## Limitations

Of this cohort, nine (39%) did not wear HAs and a large proportion had their CI evaluation performed with clinic-stock HAs (78%). Most had contralateral PTAs and WRS scores that would warrant referral for CI evaluation. Given the small sample size with strong representation of CI candidates, this represents a biased sample that could overestimate the usefulness of the SII. However, using the 40% best-aided CNC cutoff, approximately half of the ears tested were not CI candidate ears. Nonetheless, a larger study with a more heterogeneous sample should be performed. This would include patients with steeply sloping audiograms that fit potential EAS candidacy consideration where it is unknown how the SII could accurately capture this specific subpopulation.

The SII-C and SII-P were recorded with different targets and used different stimuli for calculation, which could affect the internal validity of the study by impacting SII values. Specifically, SII-C was calculated using speech at 60 dB A with NAL-N2 targets, and SII-P was calculated using pink noise at 65 dB A with NAL-RP targets. No study directly compares NAL-NL2 and NAL-RP targets on speech intelligibility. NAL-RP was specifically developed for patients with severe and profound hearing loss to maximize speech intelligibility (Mueller 2005). NAL-N2 was developed as an updated version of NAL-NL1 after empirical data necessitated adjustments to the theoretically derived formula but with the same goal of maximizing speech intelligibility (Keidser et al. 2012). NAL-NL1 prescriptions were found to be similar to those of NAL-RP at average speech input levels (Byrne et al. 2001). A future study comparing NAL-NL2 and NAL-RP prescriptions can elucidate how these targets affect SII, which may affect this and future study’s findings.

## Conclusion

The SII is an easily accessible metric that can be used to identify patients that would qualify for a CI. This pilot study has shown that the SII is able to capture at least 80% of patients who should undergo CI evaluation even under varying clinic-specific candidacy stringency (e.g., best-aided CNC ≤60%, ≤50%, or ≤40% words correct). The SII was also shown to add additional predictive value beyond traditional “60/60” referral guidelines when using logistic regression modeling. Additionally, a key finding of this study was that the SII recorded from patients’ own devices was not significantly different from the SII recorded from clinic-stock devices demonstrating its usefulness in a clinical context.

## Supporting information

Supplemental Content

## Data Availability

All data produced in the present study are available upon reasonable request to the authors

## Acknowledgements

JDN is a recipient of NIDCD training grant (T32 DC020141).

