## Supplemental Content for "Speech Intelligibility Index (SII) as a Potential Referral Metric for Adult Cochlear Implant Candidacy Evaluation"

Appendix 1: S**tandard logistic model performance and comparison of area under the receiver operating characteristic curve (AUC) and likelihood ratio test (LRT) results with word recognition scores (WRS), pure tone average (PTA), and Speech Intelligibility Index-P (SII-P) for different consonant-nucleus-consonant (CNC) score candidacy cutoffs: (a) 60% CNC cutoff, (b) 50% CNC cutoff, and (c) 40% CNC cutoff. Matched-row analyses were used for nested model comparisons. Not all candidates wore hearing aids, resulting in a reduced sample (N = 14 participants; 28 ears). WRS data were unavailable for two participants.**

| (a) Standard logistic regression model for 60% CNC cutoff | | | | | |
| --- | --- | --- | --- | --- | --- |
| Predictors | N | AUC | $\Delta$AUC | LRT $\chi$^2^(1) | p-value |
| WRS | 26 | 0.80 | - | - | - |
| WRS + SII-P | 26 | 0.78 | -0.02 | 0.2 | 0.654 |
| PTA | 28 | 0.85 | - | - | - |
| PTA + SII-P | 28 | 0.84 | -0.01 | 0.0 | 0.100 |
| WRS + PTA | 26 | 0.84 | - | - | - |
| WRS + PTA + SII-P | 26 | 0.83 | -0.01 | 0.2 | 0.665 |

| (b) Standard logistic regression model for 50% CNC cutoff | | | | | |
| --- | --- | --- | --- | --- | --- |
| Predictors | N | AUC | $\Delta$AUC | LRT $\chi$^2^(1) | p-value |
| WRS | 26 | 0.85 | - | - | - |
| WRS + SII-P | 26 | 0.89 | +0.04 | 2.1 | 0.147 |
| PTA | 28 | 0.74 | - | - | - |
| PTA + SII-P | 28 | 0.73 | -0.01 | 0.0 | 0.922 |
| WRS + PTA | 26 | 0.86 | - | - | - |
| WRS + PTA + SII-P | 26 | 0.90 | +0.04 | 3.0 | 0.073 |

| (c) Standard logistic regression model for 40% CNC cutoff | | | | | |
| --- | --- | --- | --- | --- | --- |
| Predictors | N | AUC | $\Delta$AUC | LRT $\chi$^2^(1) | p-value |
| WRS | 26 | 0.85 | - | - | - |
| WRS + SII-P | 26 | 0.89 | +0.04 | 0.0 | 0.984 |
| PTA | 28 | 0.74 | - | - | - |
| PTA + SII-P | 28 | 0.73 | -0.01 | 0.8 | 0.368 |
| WRS + PTA | 26 | 0.86 | - | - | - |
| WRS + PTA + SII-P | 26 | 0.9 | +0.04 | 0.0 | 0.854 |

CNC = consonant–nucleus–consonant, WRS = word recognition score, PTA = pure tone average, SII-P = Speech Intelligibility Index from Personal/Research, AUC = area under curve, LRT = likelihood ratio test

Appendix 2: **Mixed-effects model performance and comparison of area under the receiver operating characteristic curve (AUC) and likelihood ratio test (LRT) results with word recognition scores (WRS), pure tone average (PTA), and Speech Intelligibility Index-C (SII-C) for different consonant-nucleus-consonant (CNC) score candidacy cutoffs: (a) 50% CNC cutoff and (b) 40% CNC cutoff. Mixed-effects modeling for the 60% CNC cutoff did not converge and was therefore not retained.**

| (a) Mixed-effects logistic regression model for 50% CNC cutoff | | | | | |
| --- | --- | --- | --- | --- | --- |
| Predictors | N | AUC | $\Delta$AUC | LRT $\chi$^2^(1) | p-value |
| WRS | 43 | 0.88 | - | - | - |
| WRS + SII-C | 43 | 0.93 | +0.05 | 9.1 | 0.003 |
| PTA | 46 | 0.81 | - | - | - |
| PTA + SII-C | 46 | 0.87 | +0.06 | 5.7 | 0.017 |
| WRS + PTA | 43 | 0.88 | - | - | - |
| WRS + PTA + SII-C | 43 | 0.93 | +0.05 | 6.1 | 0.013 |

| (b) Mixed-effects logistic regression model for 40% CNC cutoff | | | | | |
| --- | --- | --- | --- | --- | --- |
| Predictors | N | AUC | $\Delta$AUC | LRT $\chi$^2^(1) | p-value |
| WRS | 43 | 0.82 | - | - | - |
| WRS + SII-C | 43 | 0.88 | +0.06 | 8.6 | 0.003 |
| PTA | 46 | 0.79 | - | - | - |
| PTA + SII-C | 46 | 0.86 | +0.07 | 6.0 | 0.014 |
| WRS + PTA | 43 | 0.82 | - | - | - |
| WRS + PTA + SII-C | 43 | 0.88 | +0.06 | 5.5 | 0.019 |

CNC = consonant–nucleus–consonant, WRS = word recognition score, PTA = pure tone average, SII-C = Speech Intelligibility Index from clinic, AUC = area under curve, LRT = likelihood ratio test
